# The link between vitamin D deficiency and Covid-19 in a large population

**DOI:** 10.1101/2020.09.04.20188268

**Authors:** Ariel Israel, Assi Cicurel, Ilan Feldhamer, Yosef Dror, Shmuel M Giveon, David Gillis, David Strich, Gil Lavie

## Abstract

**BACKGROUND:** Recent studies suggest a link between vitamin D deficiency and Covid-19 infection. In our population we observe major differences in Covid-19 incidence in ethnic groups and genders in each group.

**METHODS:** We carried out a population-based study among 4.6 million members of Clalit Health Services (CHS). We collected results from vitamin D tests performed between 2010 and 2019 and used weighted linear regression to assess the relationship between prevalence of vitamin D deficiency and Covid-19 incidence in 200 localities. Additionally, we matched 52,405 infected patients with 524,050 control individuals of the same sex, age, geographical region and used conditional logistic regression to assess the relationship between baseline vitamin D levels, acquisition of vitamin D supplements in the last 4 months, and positive Covid-19.

**RESULTS:** We observe a highly significant correlation between prevalence of vitamin D deficiency and Covid-19 incidence, and between female-to-male ratio for severe vitamin D deficiency and female-to-male ratio for Covid-19 incidence in localities (P<0.001). In the matched cohort, we found a significant association between low vitamin D levels and the risk of Covid-19, with the highest risk observed for severe vitamin D deficiency. A significant protective effect was observed for members who acquired liquid vitamin D formulations (drops) in the last 4 months.

**CONCLUSION:** In this large observational population study, we show a strong association between vitamin D deficiency and Covid-19 occurrence. After adjustment for baseline characteristics and prior vitamin D levels, acquisition of liquid vitamin D formulations is associated with decreased risk for Covid-19 infection.

## Introduction

SARS-Cov-2 is a new virus, which was first identified in December 2019, and has rapidly spread to a global pandemic of primarily respiratory illness designated as Coronavirus Disease 2019 (Covid-19). Covid-19 is associated with significant mortality, particularly among the aging population, raising considerable concerns for public health. Vitamin D appears to play a prominent role in the prevention of respiratory infections^1^. Recent reports found that SARS-Cov-2 infection rate is higher in countries with low vitamin D^2,3^, and prompted further research on this topic^4^. High rates of vitamin deficiency were found in the nations highly affected by the Covid-19 epidemic, and low vitamin D levels were found in patients with severe Covid-19 cases^5,6^.

In Israel, the general population has been so far relatively spared by the pandemic. In our health organization, an infection rate of 0.88% equal in the two genders has been observed in the general population. However, in two large ethnic minorities, we observe a particularly high Covid-19 incidence: 3.03% (3.5 higher) in the Jewish ultra-orthodox population, and 1.4% in Arab communities (1.6 higher). Moreover, the male-to-female ratio for incidence is very different in these two latter groups. In Arab communities, females were significantly more affected (1:1.5), while in ultra-orthodox communities, males were more affected (1.25:1).

Both Arabs and ultra-orthodox subpopulations tend to live in specific geographic areas. Within each of these ethnic groups, there are significant differences in lifestyle, but people who live in the same locality tend to follow a similar lifestyle, often have common ethnic origins, and more importantly, individuals tend to wear a traditional (gender-specific) attire, with more body surface covered than the general population. These could affect the ability of the body to absorb sunlight and produce vitamin D. Previous research has shown that vitamin D deficiency is much more prevalent in these two minorities, and severe vitamin D deficiency is endemic among Arab women^7^.

If there is indeed an association between vitamin D deficiency and higher rates of Covid-19 infection, then we would expect to observe a significant statistical association between the prevalence of vitamin D deficiency and Covid-19 incidence across localities. Moreover, we would expect to see similar gender-specificity for vitamin D deficiency and Covid-19 cases.

Clalit Health Services (CHS) provides comprehensive health services to over 4.6 million members, and centrally manages electronic health records (EHR) with longitudinal records for over two decades, including laboratory tests, diagnoses, and purchase of medications^8^. This provides a unique opportunity to study the association between vitamin D levels and Covid-19 incidence, as well as the impact of purchase of vitamin D supplements on the risk of Covid-19.

## Methods

### Study population and data collection

We collected from the CHS data warehouse selected variables from the EHR of patients who underwent vitamin D testing between January 1^st^ 2010, and December 31^st^ 2019. In addition to the last vitamin D level, we collected data regarding age, gender, Adjusted Clinical Group (ACG)-based comorbidity measure^9^ and the primary care clinic of the patient, as of February 2020, prior to the first Covid-19 case. The primary care clinic was used to associate a geographic region, one of the three main ethnic groups (general, ultra-orthodox, and Arab), and a 3-level socio-economic status.

We collected similar data from patients who had a positive RT-PCR test for SARS-CoV-2 since the disease outbreak until August 31^st^ 2020, with the date of the first positive test taken as index date. As controls, for each SARS-CoV-2 positive patient, we matched 10 individuals of the same age, gender, geographic region, and ACG comorbidity score, assigned the same index date, and collected EHR data in the same manner.

This study has been approved by the CHS Institutional Review Board (IRB) with a waiver of informed consent, approval number: COM-0046-20.

Patients’ data were extracted and processed from CHS data-warehouse using programs developed by the first author in Python and SQL, all identifying patient data were removed prior to the statistical analyses in accordance to the protocol approved by the CHS IRB.

### Statistical analysis

In descriptive tables, statistical significance of differences observed between groups was assessed by the Chi-Square test for categorical variables, and two-tailed T-test for continuous variables.

Incidence of SARS-CoV-2 of the disease in each locality was calculated by taking the ratio between individuals registered in the locality with a positive test, and the number of CHS members registered in this locality. Female-to-male ratio for incidence was obtained by calculating the ratio between female incidence and the male incidence in each locality.

Prevalence of severe vitamin D deficiency by locality was calculated by taking the ratio between the number of individuals of the locality for which the last measured vitamin D levels was below 30 nmol/L and the number of CHS members registered in the locality who were tested for vitamin D. All vitamin D measures were taken between years 2010-2019. Female-to-male ratio for prevalence was obtained by calculating the ratio between female prevalence and male prevalence.

We used weighted linear regressions to calculate the slope and significance of associations between severe vitamin D deficiency prevalence and SARS-CoV-2 incidence, and the slope and significance of associations between female-to-male ratio for severe vitamin D deficiency and SARS-CoV-2 incidence. Regression models were weighted by the number of positive cases in each locality. We incorporated in this analysis all localities in which at least 25 individuals of a given ethnic group had a positive RT-PCR test for SARS-CoV-2 (202 localities).

Conditional logistic regression models were fitted for estimating the odds-ratio (OR) and corresponding 95% confidence interval (CI) for the risk of a positive test for SARS-CoV-2 in individuals from the matched cohort. We assessed the odds for SARS-CoV-2 infection according to vitamin D ranges in a univariable model. We also fit several multivariable logistic models, accounting for ethnic group and acquisition of vitamin D formulations in the whole matched cohort, and in subgroups of baseline vitamin D ranges.

P-values below 0.05 were considered significant. Statistical analyses and graphs were performed using R statistical software version 3.6 (R Foundation for statistical computing).

## Results

From the beginning of the outbreak and until August 31^st^, 2020, 52,537 distinct CHS members had positive RT-PCR tests for SARS-Cov-2. Table 1 shows the prevalence of Covid-19 infection in the three studied ethnic groups. The incidence of the disease varies widely between the sub-populations and is notably more prevalent among the Jewish ultra-orthodox and Arab populations.

**Table 1:** Demographics of positive SARS-CoV-2 positive tests among CHS ethnic groups.

| Ethnic group | General (mostly Jewish) |  |  | Arab |  |  | Ultra Orthodox |  |  |
| --- | --- | --- | --- | --- | --- | --- | --- | --- | --- |
|  | Male | Female | Both | Male | Female | Both | Male | Female | Both |
| **Number of members** | 1,518,465 | 1,602,807 | 3,121,272 | 627,652 | 621,744 | 1,249,396 | 130,679 | 129,910 | 260,589 |
| **SARS-CoV-2 positive** | 13,325 | 14,081 | 27,406 | 7,031 | 10,204 | 17,235 | 4,403 | 3,493 | 7,896 |
| **Infection rate** | 0.88% | 0.88% | 0.88% | 1.12% | 1.64% | 1.38% | 3.37% | 2.69% | 3.03% |
| **Male-to-Female ratio** | 1 : 1.00 |  |  | 1 : 1.47 |  |  | 1.25 : 1 |  |  |

Between the years 2010 and 2019, 1,359,339 distinct patients (over 30% of CHS members) had their vitamin D levels measured and these records were kept in CHS databases. Results are summarized in Table 2. We found that vitamin D deficiency (< 50 nmol/L), and particularly severe vitamin D deficiency (< 30 nmol/L) was much more prevalent among Ultra-orthodox and Arabs. Arab females were particularly affected by vitamin D deficiency (81.5% of the individuals tested had vitamin D levels below 50 nmol/L, and 59.1% below 30 nmol/L).

**Table 2:** Vitamin D tests performed between years 2010 and 2019 in CHS services.

| Ethnic group | General |  | Arab |  | Ultra Orthodox |  |
| --- | --- | --- | --- | --- | --- | --- |
|  | Male | Female | Male | Female | Male | Female |
| **No. of Individuals tested** | 350,161 | 656,396 | 91,195 | 167,295 | 32,157 | 47,966 |
| **Last individual vitamin D measure, median [interquartile range]** | 58.91<br>[44.50, 74.20] | 57.60<br>[41.90, 74.10] | 44.10<br>[31.00, 59.10] | 25.10<br>[16.00, 42.70] | 49.50<br>[35.30, 65.15] | 45.50<br>[31.10, 62.40] |
| **vitamin D range %** |  |  |  |  |  |  |
| < 30 nmol/L | 8.2 | 11.2 | 23.5 | 59.1 | 16.7 | 23.3 |
| 30-50 nmol/L | 25.9 | 26.2 | 37.3 | 22.4 | 34.3 | 34.0 |
| 50-75 nmol/L | 42.0 | 38.9 | 29.4 | 12.8 | 34.5 | 29.7 |
| > 75 nmol/L | 23.9 | 23.7 | 9.8 | 5.8 | 14.5 | 13.0 |

Figure 1 displays the statistical distribution of vitamin D levels in the three ethnic groups in males (upper), and females (bottom). Both the ultra-orthodox (black) and the Arab populations (green) have lower levels (P<0.001), and we observe a remarkable left tail of Arab females with extremely low levels of vitamin D, rarely seen in the other two populations.

**Figure 1:**
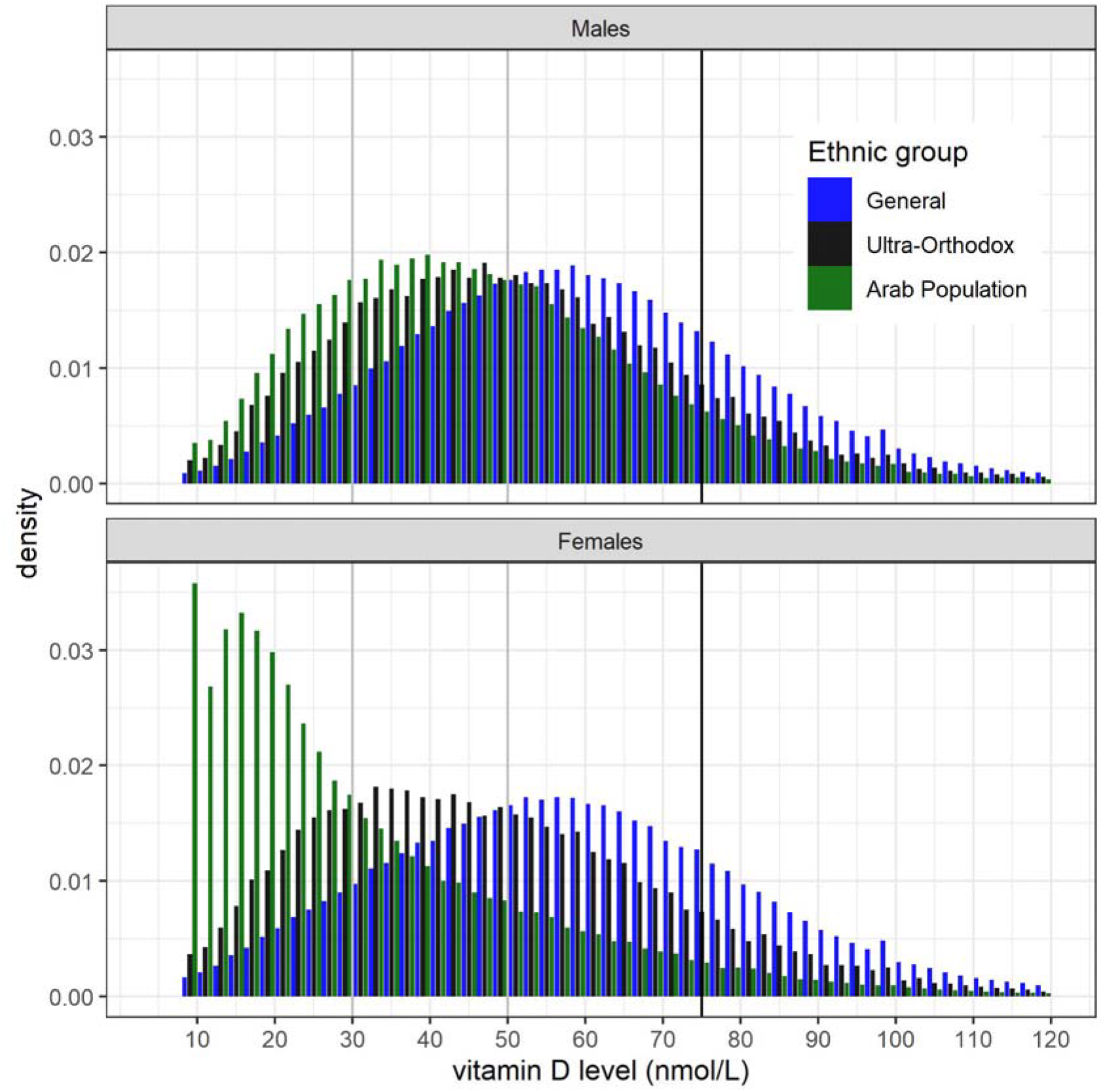
Distribution of blood vitamin D levels measured between years 2010-2020 in the three subpopulations in males (upper panel), and females (bottom panel) Histograms showing the distribution of vitamin D levels measured in males (top) and females (bottom) in each of the ethnic groups (blue for the general population, green for Arab, black for Ultra-orthodox)

Figure 2 displays the statistical distribution of baseline vitamin D levels among patients who were further tested positive for SARS-CoV-2 (red) vs. the rest of the population (grey) in males (top) and females (bottom). We found a very significant left shift of vitamin D levels, particularly striking among females (P<0.001), and a large proportion of females affected by the disease had baseline vitamin D values below 40.

**Figure 2:**
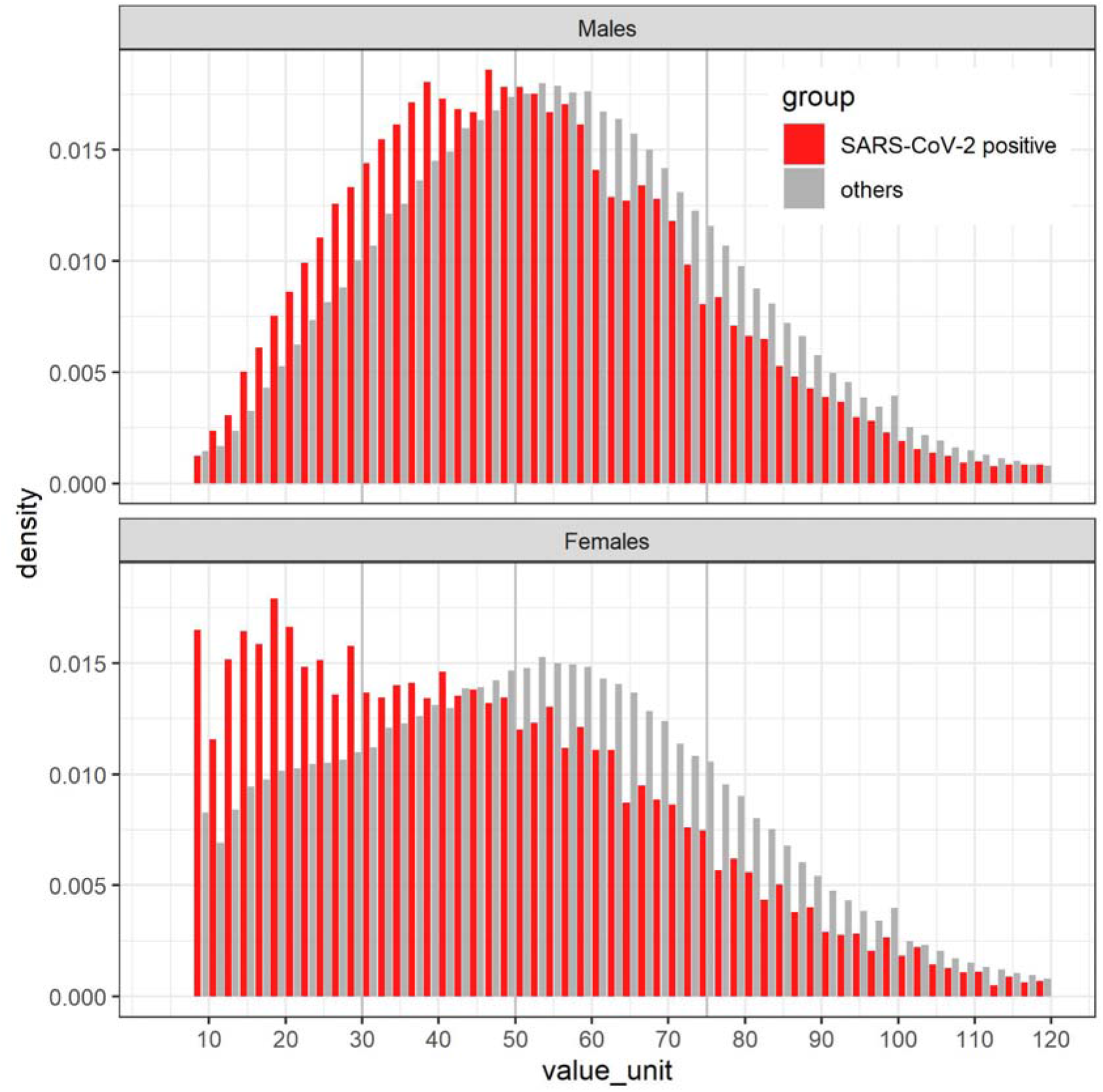
Distribution of vitamin D measured in the blood between years 2010-2020 among individuals later infected with SARS-CoV-2 patients and the rest of the population. Histograms showing the distribution of vitamin D levels measured in males (top) and females (bottom). The red histogram is for individuals who were further tested positive for SARS-CoV-2, in grey the rest of the population

Figure 3 displays the association between the prevalence of severe vitamin D deficiency (x axis) and the incidence of SARS-CoV-2 (y axis) for each of the ethnic groups (blue for the general population, black for ultra-orthodox, green for Arab) and for each locality, with each point weighted by the number of cases detected in the locality. We observe a very significant positive correlation between these two variables, both across communities (P<0.001), and within the Arab and general communities (P = 0.005 and P<0.001 respectively, panels B and C), for the orthodox population, the small number of localities did not provide enough power for this analysis.

**Figure 3:**
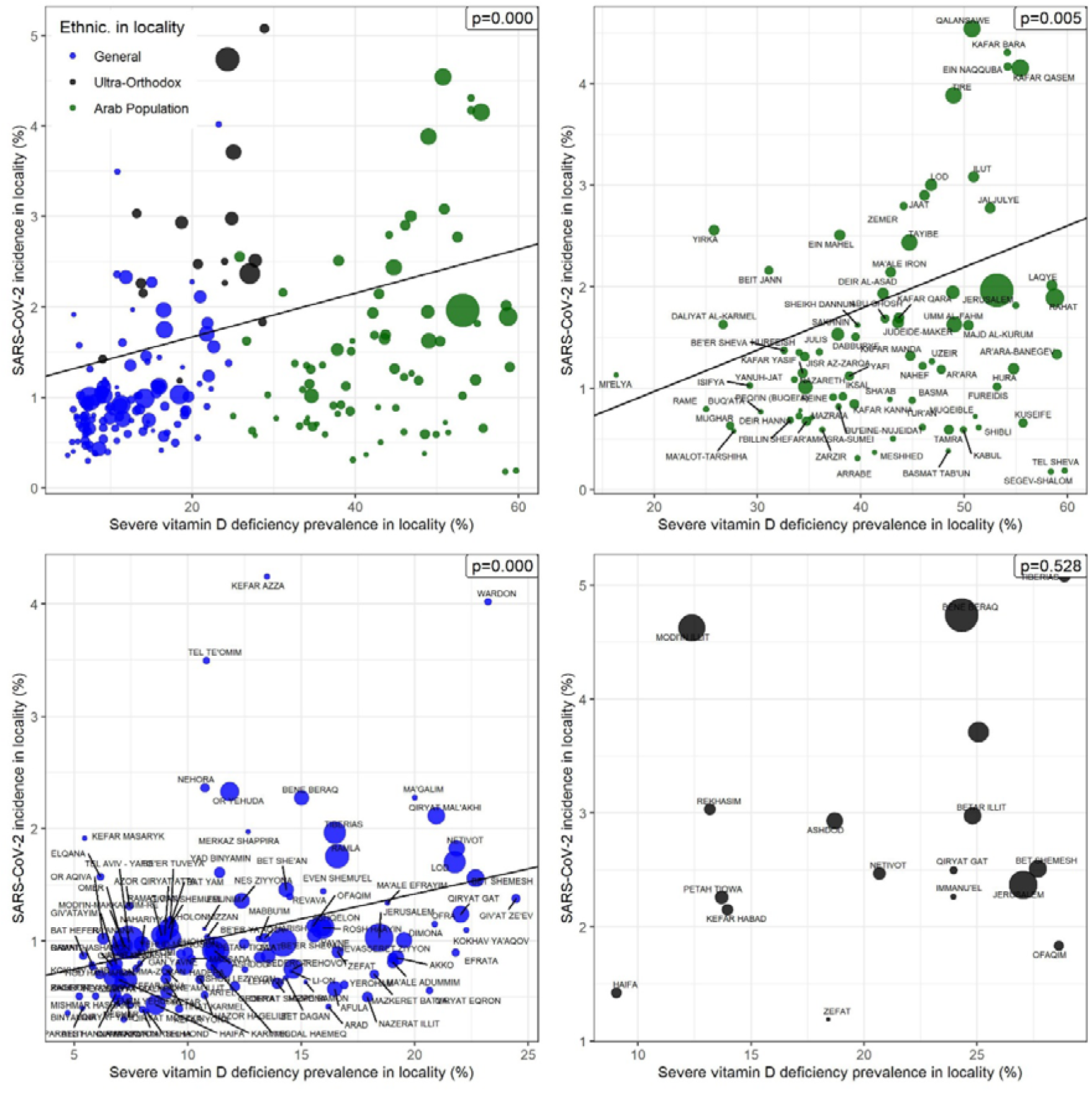
Relationship between severe vitamin D deficiency prevalence and Covid-19 incidence by ethnic group and locality. Scatter plot displaying, for each of the ethnic groups (blue for the general population, green for Arab, black for ultra-orthodox), and for each locality the relationship between the prevalence of severe vitamin D deficiency (x axis) and the incidence of SARS-CoV-2 (y axis); each spot is a locality, weights and size are proportional to the number of positive SARS-CoV-2 individuals in the locality.

Figure 4 displays in a similar manner the female-to-male ratio for severe vitamin D in each locality, and the female-to-male ratio for SARS-CoV-2 incidence. We found a highly significant positive correlation between the proportion of females affected by severe vitamin D deficiency and SARSCoV-2 incidence, both across the different groups (P<0.001), and within each group (P<0.001, P = 0.05, P = 0.007 in the Arab, general and ultra-orthodox populations respectively).

**Figure 4:**
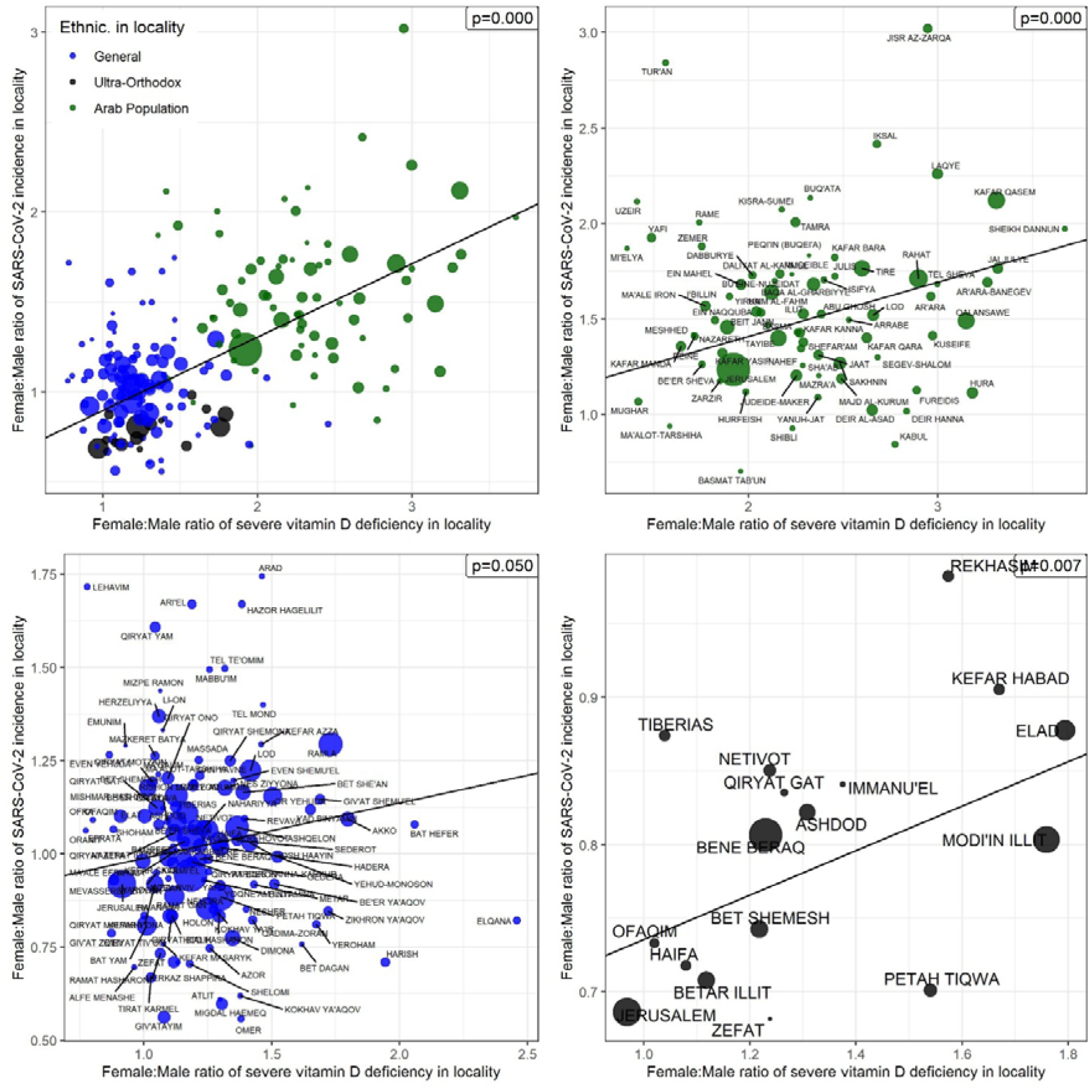
Relationship between female-to-male ratio of severe vitamin D deficiency, and female-to-male SARS-CoV-2 incidence by ethnic group and locality. Scatter plot displaying, for each of the ethnic groups (blue for the general population, green for Arab, black for Ultra-orthodox), and for each locality the relationship between female-to-male ratio for the prevalence of severe vitamin D (x axis), and the female-to-male ratio of SARS-CoV-2 incidence; each spot is a locality, weights and size are proportional to the number of positive SARSCoV-2 individuals in the locality

Having identified a strong epidemiologic correlation between severe vitamin D deficiency and SARSCoV-2 infection, we proceeded to assess, at the individual level, the effect of the different ranges of baseline vitamin D, the ethnic group, and consumption of vitamin D supplements, on the risk of SARS-CoV-2 infection. For this purpose, we used a matched cohort where each patient tested positive for the virus is matched to 10 control patients of the same age, sex, geographic region, socioeconomic status (3-level based), and the same ACG measure of comorbidity as of February 2020, prior to the onset of the pandemics. A match was found for 52,405 individuals out of 52,537 individuals who were tested positive, and 524,050 controls were assigned. The characteristics of the matched cohort are shown in Table 3.

**Table 3:** Demographics and Clinical characteristics of the cohort of patients who tested positive for COVID-19 with 10:1 matched controls.

|  | **SARS-CoV-2 cases**<br>**N=52,405** | **controls**<br>**N=524050** | **p** |
| --- | --- | --- | --- |
| **Female (%)** | 27714 (52.9) | 277140 (52.9) | 1.000 |
| **age, median** | 32.00 | 32.00 |  |
| **Interquartile range [IQR]** | [18.00, 50.00] | [18.00, 50.00] |  |
| **Region name (%)** |  |  | 1.000 |
| Jerusalem | 8931 (17.0) | 89310 (17.0) |  |
| Tel Aviv | 3161 (6.0) | 31610 (6.0) |  |
| Dan-Petach Tikva | 7819 (14.9) | 78190 (14.9) |  |
| Haifa | 5549 (10.6) | 55490 (10.6) |  |
| Center | 7588 (14.5) | 75880 (14.5) |  |
| South | 6297 (12.0) | 62970 (12.0) |  |
| Sharon-Shomron | 7778 (14.8) | 77780 (14.8) |  |
| North | 5061 (9.7) | 50610 (9.7) |  |
| Eilat | 221 (0.4) | 2210 (0.4) |  |
| **ACG comorbidity score (median [IQR])** | 0.44 [0.17, 1.67] | 0.44 [0.17, 1.67] |  |
| **Ethnic group (%)** |  |  | <0.001 |
| General | 27327 (52.1) | 350896 (67.0) |  |
| Ultra Orthodox | 7877 (15.0) | 35640 (6.8) |  |
| Arab Population | 17201 (32.8) | 137514 (26.2) |  |
| **Socio Economic Status (%)** |  |  | <0.001 |
| missing | 379 (0.7) | 2085 (0.4) |  |
| low | 27100 (51.7) | 211998 (40.5) |  |
| medium | 18095 (34.5) | 198646 (37.9) |  |
| high | 6831 (13.0) | 111321 (21.2) |  |
| **last measured vitamin D level, median [IQR]** | 45.50<br>[28.60, 63.90] | 51.20<br>[33.90, 68.50] |  |
| **vitamin D range (%)** |  |  | <0.001 |
| < 30 nmol/L | 5011 (27.3) | 34474 (20.4) |  |
| 30-50 nmol/L | 5404 (29.4) | 47042 (27.9) |  |
| 50-75 nmol/L | 5345 (29.1) | 57246 (33.9) |  |
| > 75 nmol/L | 2601 (14.2) | 30111 (17.8) |  |
| **Acquisition of vitamin D supplements 120 to 15 days before index-date (%)** | 2794 (5.3) | 25972 (5.0) | <0.001 |
| **Acquisition of vitamin D drops, 120 to 15 days before the index date (%)** | 1722 (3.3) | 17383 (3.3) | 0.714 |
| **Acquisition of vitamin D tablets, 120 to 15 days before the index date (%)** | 1004 (1.9) | 7888 (1.5) | <0.001 |

Table 4 displays the conditional logistic regression results for SARS-CoV-2 infection status in the matched cohort. Model (1) is based only on baseline vitamin D level ranges: compared to vitamin D levels above 75, which served as reference, severe vitamin D deficiency (< 30 nmol/L) carries the highest risk OR 1.817 (95% CI 1.717 to 1.924), but even the relatively high range of 50 and 75 nmol/L is associated with significantly increased risk. Model (2) is a multivariable model incorporating the ethnic group: living in an ethnic group where there is high prevalence of SARS-CoV-2 infection incurs by itself a significant risk (OR 3.442 for individuals living in Ultra-orthodox communities, and 2.618 among Arabs), but even after controlling for this factor, vitamin D levels are associated a significant increase in risk for the individuals, even for the 50–75 nmol/L range. Model (3) incorporates the purchase of vitamin D formulations 120 days to 15 days before the index date. When studying separately individual vitamin D formulations available in CHS pharmacies, we were surprised to observe diverging results, with acquisition of some vitamin D formulations associated with significantly decreased risk for SARS-CoV-2, while others were associated to significantly increased risk. Interestingly, the common feature of the vitamin D formulations which were associated with decreased risk were that they were provided as drops, so we grouped acquisition of vitamin D drops these under one variable, tablet-form being the second most common other form, we grouped their acquisition as another variable. After controlling for ethnic group and baseline vitamin D levels, acquisition of vitamin D drops was associated with a significant decrease in risk OR = 0.905 (95% CI 0.848-0.967), and acquisition of vitamin D tablets was associated with a significant increase in risk OR = 1.248 (95% CI 1.152-1.352). Models (4) (5) and (6) are subgroup analyses which study the impact of acquisition of vitamin D formulations in subgroups of patients with different ranges of baseline vitamin D (4): below 50, (5) above 50, (6) above (75). Interestingly, acquisition of vitamin D drops are associated with decreased risk in each subgroup, suggesting that liquid vitamin D supplementation could protect from SARS-CoV-2 infection in almost all individuals, regardless of its vitamin D levels.

**Table 4:** Conditional logistic regression models for estimating odds ratio for SARS-CoV-2 infection status and 95% confidence intervals based on baseline factors in matched cohort.

| Model type | Univariable | Multivariable |  |  |  |  |
| --- | --- | --- | --- | --- | --- | --- |
|  | (1) | (2) | (3) | (4) | (5) | (6) |
| Explanatory variables | vitamin D levels at baseline | Vitamin D levels at baseline and ethnic group | Vitamin D levels at baseline, ethnic group, acquisition of vitamin D | Ethnic group and acquisition of vitamin D |  |  |
| Subgroup analyzed |  |  |  | baseline vit. D below 50 nmol/L | baseline vit. D above 50 nmol/L | baseline vit. D above 75 nmol/L |
| **Baseline vitamin D range** |  |  |  |  |  |  |
| < 30 nmol/L | 1.817<br>(1.717, 1.924)<br>p = 0.000 | 1.275<br>(1.199, 1.355)<br>p = 0.000 | 1.27<br>(1.195, 1.351)<br>p = 0.000 |  |  |  |
| 30-50 nmol/L | 1.37<br>(1.297, 1.446)<br>p = 0.000 | 1.186<br>(1.122, 1.254)<br>p = 0.000 | 1.183<br>(1.118, 1.251)<br>p = 0.000 |  |  |  |
| 50-75 nmol/L | 1.097<br>(1.039, 1.158)<br>p = 0.001 | 1.057<br>(1.001, 1.117)<br>p = 0.047 | 1.053<br>(0.997, 1.113)<br>p = 0.064 |  |  |  |
| >75 nmol/L | ref. | ref. | ref. |  |  |  |
| **Ethnic group** |  |  |  |  |  |  |
| General population |  | ref. | ref. | ref. | ref. | ref. |
| Ultra-orthodox |  | 3.442<br>(3.251, 3.644)<br>p = 0.000 | 3.445<br>(3.254, 3.647)<br>p = 0.000 | 3.415<br>(3.133, 3.722)<br>p = 0.000 | 3.52<br>(3.163, 3.917)<br>p = 0.000 | 4.631<br>(3.416, 6.279)<br>p = 0.000 |
| Arab |  | 2.618<br>(2.488, 2.755)<br>p = 0.000 | 2.618<br>(2.488, 2.755)<br>p = 0.000 | 2.625<br>(2.448, 2.815)<br>p = 0.000 | 3.013<br>(2.710, 3.349)<br>p = 0.000 | 3.84<br>(2.902, 5.081)<br>p = 0.000 |
| Acquisition of vitamin D drops 120 to 15 days before the index date |  |  | 0.905<br>(0.848, 0.967)<br>p = 0.004 | 0.899<br>(0.805, 1.005)<br>p = 0.063 | 0.889<br>(0.808, 0.977)<br>p = 0.016 | 0.81<br>(0.673, 0.975)<br>p = 0.027 |
| Acquisition of vitamin D tablets 120 to 15 days before the index date |  |  | 1.248<br>(1.152, 1.352)<br>p = 0.00000 | 1.184<br>(1.037, 1.352)<br>p = 0.013 | 1.218<br>(1.080, 1.373)<br>p = 0.002 | 1.124<br>(0.874, 1.446)<br>p = 0.362 |
| No. Observations | 187,234 | 187,234 | 187,234 | 91,515 | 95,719 | 32,986 |

## Discussion

In this large population study on individuals of diverse ethnic groups, we have uncovered what appears to be a strong and significant association between low vitamin D levels and the risk of SARSCoV-2 infection. Individuals with low baseline vitamin D levels were significantly more prone to get infected with SARS-CoV-2. Moreover, marked variations in infection rates were observed in the different studied communities, and they appear to largely reflect the pattern of vitamin D deficiency within these communities. The highest risk being observed among individuals with severe vitamin D deficiency living in communities where many individuals have low vitamin D. Conversely, individuals living in communities with a low rate of severe vitamin D deficiency seem to benefit from a “heard immunity” effect, probably because their neighbors are less likely to spread the virus to them.

To the best our knowledge, this is the first study to show such a profound and significant association between vitamin D deficiency and SARS-CoV-2 infection rate. Several recent studies showed that northerly latitude is associated with higher mortality rate and hospitalization rate for Covid-19 worldwide^2^. Several potential mechanisms have been proposed to explain the observed association between vitamin D levels and the risk of Covid-19 infection^10^. Notably, viruses could disrupt the cell junction integrity^11^, while vitamin D may maintain cell junctions and hence decrease the risk of infection; vitamin D also enhances cellular innate immunity partly through the induction of antimicrobial peptides which can interfere with viral replication^12^.

In our study, we observe that vitamin D supplementation, particularly in the form of drops, provides a significant protection against SARS-CoV-2 infection. To our knowledge, this is the first population study to identify a significant protective effect for vitamin D formulations against SARS-CoV-2. The ability to account for baseline vitamin D levels, as well as the matched cohort design, allowed us to overcome the potential confounding effects of other factors such as age, gender, socioeconomic status, previous comorbidity and geographic region.

We acknowledge our study’s limitations as being observational, noting the difficulty in eliminating all possible confounders. Notably, vitamin D supplements being available “over the counter” in pharmacies and stores, an unknown number of CHS members might have purchased vitamin D supplements with no trace in our electronic records, so our study might not have apprehended the full effects of vitamin D supplementation.

Besides the link between vitamin D levels, vitamin D acquisition, and SARS-CoV-2 infection rate, our study made two intriguing observations that deserve attention. First, in our regression models, vitamin D drops were associated with decreased risk for SARS-CoV-2, but vitamin D tablet formulations were associated to increased risk. In addition, males from ultra-orthodox communities tend to have higher rates of SARS-CoV-2 infection than females, even though their measured vitamin D levels at baseline were generally not lower than in females. We propose a putative explanation for these observations: the virus port of entry is the oropharynx, it is where it first reaches mucosal membranes, initially replicates and causes its first detectable effects (anosmia, agusia, sore throat). High vitamin D concentration in the oropharynx might be the most important factor that prevents this initial infection and replication. Vitamin D in drop forms is likely mostly absorbed by the mucous membranes of the oropharynx, and the vitamin D concentration there is likely to be elevated following drops intake. Conversely, vitamin D tablets are absorbed further in the gastrointestinal track, and vitamin D concentration reaching back the oropharynx might not provide adequate protection. In addition, it is likely that vitamin D tablets acquisition is a confounder for low vitamin D levels, as individuals who purchase these (slightly more expensive) vitamin D supplements do so because they know their vitamin D levels to be low; therefore, vitamin D tablet acquisition might be a marker of low vitamin D levels, without the benefit of vitamin D drops increasing oropharyngeal concentration. Patients who purchased vitamin D tablets are also not likely to have acquired vitamin D drops in a non CHS managed pharmacy, as opposed to other people for which we have no trace of vitamin acquisition. Moreover, taking tablets out of their package requires digital manipulation of an object that is orally ingested. If the patient did not sterilize his hands before this procedure, this might be the exact way by which the virus gets inside the oropharynx where it could replicate; taking drops directly from the bottle does not involve such a risk. As for the high incidence among ultra-orthodox males: first, we notice that religious men frequently have beards, they also wear hats with large borders that shadow the face from sunlight. Both beards and hats are large surfaces in direct contact to the face, where the virus could deposit until a hand provides the contact with mucous membranes. In addition, the shadow of the hat and of the beard probably prevents vitamin D from being synthesized in the skin of the face, in proximity to the oropharynx where it could prevent initial virus replication. These would explain why at equal blood levels of vitamin D, ultra-orthodox men tend to get more infected than women. Alarming infection rates in Ultra-orthodox Jewish communities relatively to the general population have been reported in other countries in Europe and in America as well^13^.

The vitamin D hypothesis also provides attractive explanations for many of the observations that were made so far regarding the epidemiology of Covid-19. We^14^, like others, have found a significantly decreased infection rate among smokers. Smoking being prohibited in Israel in most workplaces and public buildings, smokers are much more inclined to spend time outdoors throughout the day in order to smoke, and therefore probably get more sunlight exposure than nonsmokers. In addition, we and others^14^ found that overweight individuals and individuals with hypertension – a condition closely associated with high body mass index – have significantly higher rates of SARS-CoV-2 infection and were more likely to suffer from complications of the disease^15^. Vitamin D is generally low in overweight individuals^16^: being lipid soluble^17^, vitamin D is rapidly taken up by adipose tissues, and a smaller dose might reach the oropharynx where it could provide protection from the virus.

There is also an enigma: how comes, given the proven capacity of this coronavirus to mutate and spread at a very high rate among humans, that humans and other mammals were apparently relatively spared from coronaviruses until the current pandemics. How is it that among all mammals, bats are the principal reservoir of hundreds of coronaviruses strains? Interestingly, bats live mostly in the dark, and their vitamin D levels are so low that they are often undetectable^18^. Bats have developed alternative pathways to regulate bone mineralization^19^, but the lack of vitamin D associated with living in the darkness, might impair bats from eliminating these viruses, and this may explain why so many coronaviruses are present in bats^20^. Naturally, almost all other mammals live in the free air and get abundant sunlight, and even humans, throughout history, had to spend a large part of the day outdoors in order to hunt, produce and gather food. Only the technological advances of modern times have enabled humans to live and prosper while staying in the confines of acclimatized buildings, behind windows protected by ultraviolet filters, enlightened from artificial light sources. These could be the environmental factors that finally enabled the rapid spread of this virus strain among humans.

It is remarkable that the current pandemic began in December in China, spread rapidly to countries of the Northern hemisphere in the midst of the winter and that the first wave began its downslope during the spring, when days became progressively longer, while at the same time spreading rapidly in countries of the southern hemisphere, where days were becoming shorter. Most of the African continent, where sun is abundant and people wear light garments, appears to have been spared by this pandemic. Since June 21, days begin to shorten again in the North, and a second wave is currently observed, including in Israel. The hypothesis of a natural protection provided by sunlight-enabled vitamin D provides a possible explanation for these observations. Naturally, additional human factors not related to sunlight might have contributed to the wave pattern.

A similar seasonal pattern is in fact observed for most respiratory viruses, and influenza in particular. Why influenza propagates almost exclusively in the winter despite the fact that humans live in closed spaces all over the year is unclear. Vitamin D might be the culprit. Vitamin D synthesized with sunlight provides a natural protection against influenza and respiratory viruses^21,22^. This protection is lacking in the winter when days are short. Indeed, a study performed in our health organization showed a significant seasonal variation in vitamin D levels^7^.

Naturally, these hypotheses need to be confirmed in further studies, but we believe that our findings deserve attention. Our observations might guide policymakers to adopt interventions that are effective against this virus, before the second wave amplifies and increases the death toll. In this context, some policies might deserve reevaluation, such as confinement of individuals in closed buildings, and wearing outdoors a facemask that prevents sunlight from reaching the face and oropharynx area.

## Conclusion

Results from this study suggest that populations should be urged to get more sunlight exposure in order to decrease Covid-19 risk. Oral vitamin D uptake should be encouraged, preferably in the form of drops.

## Data Availability

Access to data is restricted

## Acknowledgement

All authors have no conflict of interest to report

